# Sufficient COVID-19 quarantine and testing on international travelers from China

**DOI:** 10.1101/2023.11.03.23297426

**Authors:** Dinesh Bojja, Scott Zuo, Jeffrey P. Townsend

**Affiliations:** Yale College, New Haven, Connecticut 06520, USA; Department of Biostatistics, Yale School of Public Health, New Haven, Connecticut 06510, USA; Program in Microbiology, Yale University, New Haven, Connecticut 06511, USA; Program in Computational Biology and Bioinformatics, Yale University, New Haven, Connecticut 06511, USA

**Author notes:** ***Corresponding Author*** *Jeffrey P. Townsend PhD*, 135 College St, New Haven, CT 06510-2483.

**Keywords:** Quarantine, Pandemics, Prevalence, COVID-19, World Health Organization, China, Policy

## Abstract

**Objectives:** Removal of zero-COVID restrictions in China led to a surge in COVID-19 cases. In response, countries imposed restrictions on Chinese travelers. However, border policies may not provide substantial benefits and their assessment depends on accurate prevalence data.

**Methods:** We analyzed quarantines and testing sufficient to prevent additional in-country transmission for February 13–19, 2023 based on World Health Organization (WHO) and self-reported infection rates to estimate prevalence.

**Results:** Here we have shown that self-reported prevalence data indicated more stringent border restrictions compared to WHO-published prevalence statistics. No travel restrictions were required for Singapore for infections to not be greater than in complete border closure, while a 1-day quarantine, 2-day quarantine, and a 3-day quarantine were indicated for England, Germany, and Scotland respectively. A 10-day quarantine, 11-day quarantine, and 13-day quarantine were required for Italy, Japan, and France, respectively, to prevent an increase in the number of within-country infections due to travel, while South Korea required a complete border shutdown.

**Conclusions:** Our results demonstrated the necessity for accurate and timely reporting of pandemic statistics to prevent an increase in viral spread. Through the minimum-quarantine analysis, countries can use science to determine policy, minimize international friction, and improve the cost-efficiency of interventions.

## Introduction

Early in the pandemic, the government of China imposed strict “zero-COVID” lockdown measures that successfully prevented the spread of the SARS-CoV-2 virus throughout its populations (*1*). However, this policy required of the populace a substantial socioeconomic tradeoff. In response to widespread protests in December 2022, the Chinese government suddenly lifted its restrictions (*2, 3*). The largely unvaccinated Chinese population was subjected to a rapid increase in cases, with infection and death rates skyrocketing over December and January (*4*).

Fearing that travelers from China would spur yet another wave of COVID, other countries imposed restrictions on Chinese travelers to prevent an influx of the virus, even though ongoing community transmission was ubiquitous elsewhere. It has been long understood that border controls can help stop disease transfer by localizing the virus and protecting those who are not yet infected (*5*). At the start of the pandemic, border controls instituted around the world effectively delayed viral spread (*6, 7*). However, with near-worldwide ongoing community transmission by 2022, it was no longer clear that international border controls substantially decreased infection within countries (*8, 9*).

Here we compiled statistics regarding national population demographics, COVID rates, and international travel rates (**Supplementary Materials 1**). We extracted demographic data, travel data, COVID vaccination uptake, and infection prevalences from national and World Health Organization (WHO) databases and compared them to distinct COVID prevalence estimates based on self-reported infection rates of Chinese citizens (*10*). Based on these data, we estimated quarantine durations for each destination country that were sufficient to prevent an increase in infections when compared to a complete border closure (*8*).

## Methods and data collection

Population sizes for all countries were based on country-specific census data. Travel data between China and each of the other destination countries were based on travel during pre-pandemic times scaled to predictions of total travel to and from China in 2023, or from late 2022 travel (data utilized and sources for each can be found in the **Supplementary Materials**). COVID prevalence data for February 13–19, 2023 were derived from weekly infection rates provided by the WHO COVID Dashboard. For China, the prevalence of COVID-19 was also alternatively estimated based on the self-reported infection rate of Chinese citizens from February 2–4, 2023 (*10*).

For the vaccination status to be considered efficacious during the span of our analysis, we collected and applied the rate of vaccination in each country only for individuals receiving it between mid-2021 and February 2023. Vaccinations received in China are mostly inactivated viral vaccines (*11*). Accordingly, we used 20.5 months based on the Ad26.COV2.S inactivated adenovirus vaccination (*12*) for the durability of immunity for those vaccinated in China. We specified 21.5 months for the durability of immunity from natural infection (*12*): individuals tallied within the cumulative infections reported by the WHO COVID Dashboard between mid-2021 and February 2023 were considered immune in the study. These data were supplied to a model that computed the destination country quarantine and testing approach sufficient to match or better the in-country transmission expected from complete border closure (*8*).

### Sufficient quarantines by test type and country

To determine the suggested quarantine, the number of imminent infections in the destination country at each duration of quarantine was calculated under each type of testing and is compared to the number of imminent infections with no travel. The quarantine duration where the imminent infections are equal was considered the minimum sufficient quarantine. The minimum sufficient quarantine tended to stay relatively constant across the different testing methods in each nation (**Tables 1–2**). Of the analyzed testing methods, RT-PCR testing allowed countries to minimize their quarantine duration, making it the most preferred testing method for effective border controls.

**Table 1:** Minimum sufficient days of quarantine based on World Health Organization Data.

| Country | No Test | RT-PCR | Rapid antigen exit | Rapid antigen entry and exit |
| --- | --- | --- | --- | --- |
| Scotland | 1 | 0 | 0 | 0 |
| England | 0 | 0 | 0 | 0 |
| France | 12 | 12 | 12 | 12 |
| Germany | 0 | 0 | 0 | 0 |
| Italy | 9 | 9 | 9 | 9 |
| Singapore | 0 | 0 | 0 | 0 |
| Japan | 11 | 11 | 11 | 11 |
| South Korea | >14 | >14 | >14 | >14 |

**Table 2:** Minimum sufficient days of quarantine based on self-reported survey.

| Country | No Test | RT-PCR | Rapid antigen exit | Rapid antigen entry and exit |
| --- | --- | --- | --- | --- |
| Scotland | 7 | 3 | 5 | 3 |
| England | 3 | 1 | 1 | 1 |
| France | 13 | 13 | 13 | 13 |
| Germany | 3 | 2 | 2 | 1 |
| Italy | 11 | 10 | 11 | 10 |
| Singapore | 2 | 0 | 0 | 0 |
| Japan | 11 | 11 | 11 | 11 |
| South Korea | >14 | >14 | >14 | >14 |

Since RT-PCR testing is generally considered to be the most rigorous and self-reported survey data for prevalence in China is generally considered a more reliable metric for the prevalence of infection in China, we here focus on trends in the sufficient quarantines with RT-PCR testing based on self-reported survey data for prevalence in China. Sufficient quarantines for Chinese travelers in each destination country varied significantly depending on the source of the prevalence data. Quarantines for countries with fewer daily inbound travelers from China tended to have shorter minimum durations to be sufficient to match complete border closure. For example, the suggested quarantine for Scotland, with 226 travelers daily, was only 3 days in duration using RT-PCR testing; no quarantine using the WHO data for prevalence in China (**Figure 1A–B**). With fewer inbound travelers, a less stringent quarantine was required to prevent an increase in imminent infections. Countries with higher immunity—whether infection-derived or vaccine-derived—tended to require stricter quarantines to be sufficient to match border closure. Sufficient quarantines for France and South Korea, were 13 days and >14 days, respectively; 12 days and >14 days, respectively, using WHO data for prevalence in China (**Figure 1E–F, Figure 2E–F**). In countries with high immunity to COVID-19, inbound tourists from China with infection would add a high number of imminent infections—more infections than the number of in-country infections under complete border closure. For the rest of the countries in the analysis, these trends remained consistent (**Figs. 1–2**).

**Figure 1:**
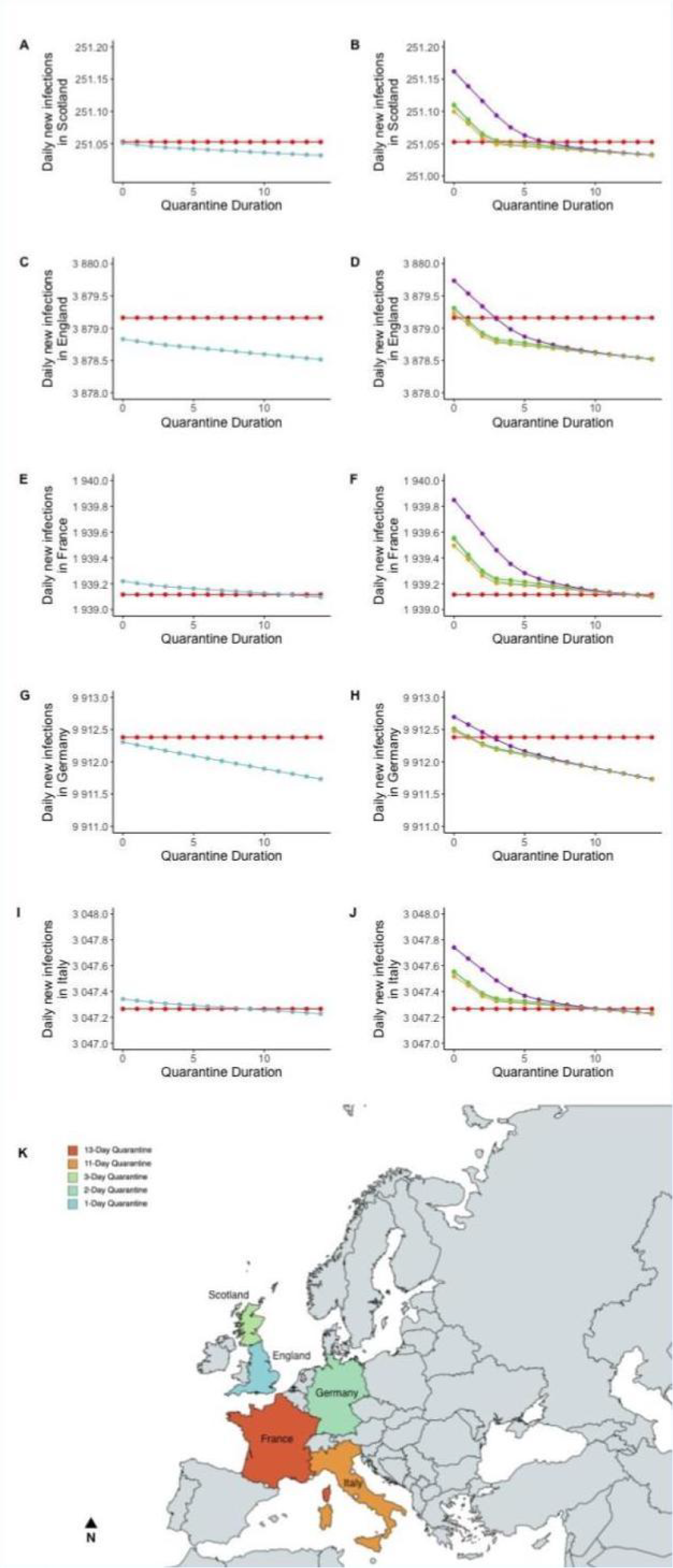
Recommended travel quarantines for European countries. Recommended travel quarantines for Scotland based on Chinese prevalence data from (**A**) World Health Organization data from China and (**B**) self-reported infection rates from Chinese citizens; for England based on (**C**) WHO data and (**D**) self-reported rates, for France based on (**E**) WHO data and (**F**) self-reported rates, for Germany based on (**G**) WHO data and (**H**) self-reported rates, and for Italy based on (**I**) WHO data and (**J**) self-reported rates. Differences in daily new infections among travel restrictions are negligible based on (**A, C, E, G**, and **I**) World Health Organization data from China, but are more substantial based on the self-reported infection rates (**B, D, F, H**, and **J**), where policies of no testing (purple), RT-PCR vs isolation test (blue) a rapid antigen quarantine exit vs isolation test (green), a rapid antigen quarantine entry and exit vs isolation test (yellow), and a complete travel ban (red). Sufficient minimum durations of quarantines with RT-PCR testing to ensure that in-country transmission will not increase due to travel compared to a complete travel ban, based on Chinese self-reported infection rates [10], are mapped to (**K**) Scotland (Green, 3-Day Quarantine), England (Blue, 1-Day Quarantine), France (Reddish Orange, 13-Day Quarantine), Germany (Aquamarine, 2-Day Quarantine), and Italy (Orange, 10-Day Quarantine).

**Figure 2:**
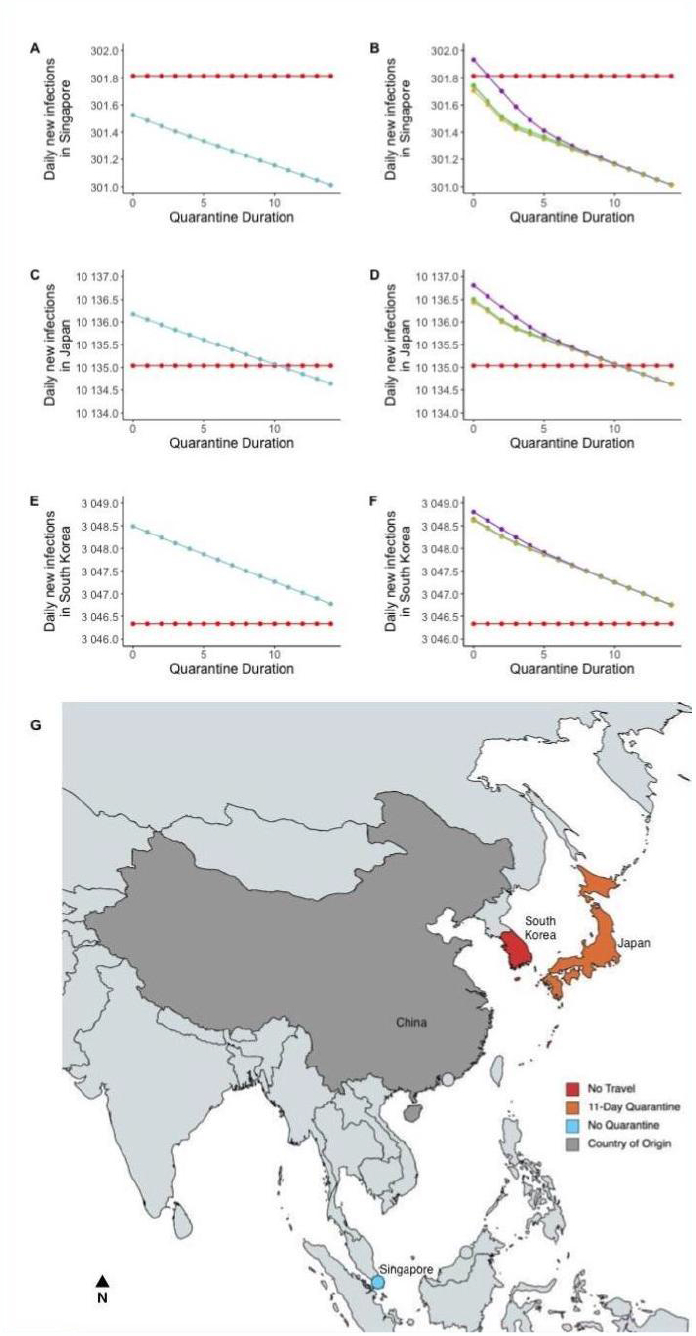
Recommended travel quarantines for Asian countries. Recommended travel quarantines for Singapore based on Chinese prevalence data from (**A**) World Health Organization data from China and (**B**) self-reported infection rates from Chinese citizens; for Japan based on (**C**) WHO data and (**D**) self-reported rates, and for South Korea based on (**E**) WHO data and (**F**) self-reported rates. Differences in daily new infections among travel restrictions are negligible based on (**A, C, E**) World Health Organization data from China, but are more substantial based on the self-reported infection rates (**B, D, F**), where policies of no testing (purple), RT-PCR vs isolation test (blue) a rapid antigen quarantine exit vs isolation test (green), a rapid antigen quarantine entry and exit vs isolation test (yellow), and a complete travel ban (red). Sufficient minimum durations of quarantines with RT-PCR testing to ensure that in-country transmission will not increase due to travel compared to a complete travel ban, based on Chinese self-reported infection rates [10], are mapped to (**G**) Singapore (Blue, No Quarantine), Japan (Reddish Orange, 11-Day Quarantine), and South Korea (Red, No Travel).

The strategy of travel restriction sufficient to enable travel without increasing within-country transmission as well as the recommended length of quarantine could differ significantly for different countries in our analysis. Some factors that influenced these differences include the duration of travel and the number of travelers, as increased travel would require more stringent restrictions to prevent excessive disease transfer. Differing vaccine coverages also enable nations to have more relaxed border controls, as individuals are well-protected from foreign infection. Other factors, such as disease prevalence and population size, also have an effect on the sufficient quarantine.

### Implications for COVID-19 policy

Here we have shown that using WHO data from February 2023, a quarantine longer than 14 days against Chinese travelers was required to prevent an increase in infection in South Korea. A minimum sufficient quarantine duration of 9 days, 11 days, and 12 days was required for Italy, Japan, and France to prevent an increase in in-country infection due to travel respectively. No quarantine was necessary to prevent an increase in infections when compared to complete border closure for Scotland, England, Germany, and Singapore. Alternatively, when using self-reported infection rates from Chinese citizens, a quarantine longer than 14 days against Chinese travelers was still required to prevent an increase in infection in South Korea, while a minimum sufficient quarantine duration of 10 days, 11 days, and 13 days was required for Italy, Japan, and France to prevent an increase in in-country infection due to travel respectively. A minimum sufficient quarantine of 1 day, 2 days, and 3 days was required for England, Germany, and Scotland, respectively, while no quarantine was required for Singapore.

Our result demonstrates the importance of documenting and publishing accurate and timely information regarding COVID-19 rates in different countries. The official WHO case numbers could be used in the analysis (*13*). However, these numbers, supplied by Chinese officials, may be under-representative of the actual infection rate in China (*14*). COVID prevalence estimates in China were far lower using officially published statistics from China through the World Health Organization (*13*) than when using alternative estimates of prevalence such as the self-reported rate of infection (*10*). When comparing results using WHO data and self-reported infection rates, it is evident that an accurate as well as precise estimate of the prevalence of COVID in China is essential to policy decision-making, as the survey-based estimate leads to a substantially greater number of additional infections due to travel, and requires a longer quarantine to prevent the rate of in-country infections from increasing. When analyzing the surveyed Chinese travelers to Italy who tested positive for COVID-19, the prevalence could be estimated to be 22.7%, which would require even more stringent quarantines (*15*).

Our results illustrate how multiple additional factors can affect the transmission prevalence for traveling. Length of stay in the destination or origin country and the vaccination and infection rates in each country can each lead to substantial changes in the infection rate consequent to permitting travel. As a pandemic progresses, sufficient quarantines vary as the prevalence of disease, vaccine efficacy, and travel rates change. Countries can make beneficial decisions only when they consider up-to-date data regarding the wide range of factors with corresponding data before imposing restrictions. Governmental, non-governmental, and commercial entities can utilize these tools as well as country-specific and time-sensitive data to assist their decision-making process. Such advised decision-making will likely provide better public health benefits than responding to domestic or international political pressures or emulating other nations that have distinct circumstances with regard to disease prevalence, natural or vaccine-mediated immunity, population size, demographics, and rates of travel.

When comparing the imminent infection with different border control strategies for each nation, there is little difference between the infection rates, often within margins of difference of less than one infection per day. However, the small scale of these differences emphasizes the importance of weighing the practicality of the quarantine strategy. If there is no tangible difference between enforcing a travel restriction or not, it may be in a country’s best interest to refrain from border controls to spend the money in better ways. Rather than enforcing a quarantine, there may be better, more productive ways to spend the money, such as for case-finding and isolation of positive cases.

Numbers of inbound and outbound travelers, as well as the length of stay in each destination country, are only estimates instead of real-time data. Since the data is scaled using data from years before the pandemic, they may not be representative of traveling habits after recovery, which could misrepresent the viral spread due to travel between select two countries. Access to more accurate, fine-scaled data for recent travel and vaccination would yield more realistic results that are representative of current travel patterns. These results would be more applicable and less uncertain with regard to the efficacy of travel restrictions.

It should be cautioned that the prevalence of COVID-19 in a country may provide an inaccurate estimate of the number of people actively traveling with an infection. It is probable that individuals recently exposed to the virus, but not necessarily yet symptomatic, will self-select and choose not to travel, which reduces the number of transmissions across borders relative to the number of infections within a country. In conditions where quarantines are justified, countries can obtain the most relevant information by performing surveillance sampling of travelers from diverse origins.

## Conclusion

Quarantine durations for different countries on travelers from China were determined such that they were sufficient to prevent an increase of infection in the destination country when compared to a strategy of complete border closure. These quarantines were calculated using two different estimates for the prevalence of COVID-19 in China: official WHO data and self-reported infection rates of Chinese citizens. Compared to using the WHO data from China, using prevalence based on self-reported infection rates yielded a stricter minimum sufficient quarantine duration. Of the analyzed countries, South Korea would have had to ban travel to prevent increased within-country transmission. Italy could have imposed a 10-day quarantine, Japan could have imposed an 11-day quarantine, and France could have imposed a 13-day quarantine to prevent an increase in infections. On the other end of the spectrum, England could have imposed only a 1-day quarantine, Germany could have imposed a 2-day quarantine, and Scotland could have imposed a 3-day quarantine with RT-PCR testing, while Singapore would be sufficiently served by RT-PCR testing and no quarantine to prevent any increase in in-country infection rates.

Our analysis illustrates how modeling disease transfer and border policies can substantially and quantitatively inform future policy decisions on healthcare-oriented quarantines. Further, it demonstrates the importance for nations to publish accurate infection statistics to make informed policy decisions. This approach should be applied in the evaluation of future policy decision-making as nations weigh the public health effects of quarantine with the economic and social effects.

## Supporting information

https://zenodo.org/records/8194600

## Data Availability

All raw data, documentation, sourcing, and analysis files can be accessed via Zenodo:
https://zenodo.org/record/8194600

## Acknowledgments

Funding: This work was supported by the National Science Foundation of the United States of America (CCF 1918784 to Jeffrey P. Townsend).

## Supplemental

All raw data, documentation, sourcing, and analysis files can be accessed via Zenodo: https://zenodo.org/record/8194600

