## Supplementary material for "Sufficient COVID-19 quarantine and testing on international travelers from China": https://zenodo.org/records/8194600

### *Data Collection*

The exact documentation of the sources used to find specific data is described here. All related data can be found in ChinaCOVIDRawData.xlsx. These data are exact as of June 18th, 2023.

**China (WHO Data)**

| Data | Link and Extraction Methods |
|---|---|
| Vaccine coverage: At least one dose<br><br>ChinaWHO!B2:H2 | https://ourworldindata.org/covid-vaccinations<br><br>The coverage for individuals who took at least one dose of the vaccine during the 20.5-month period of vaccine durability was calculated by subtracting the cumulative percentage from Feb 9th, 2023, by the cumulative percentage from Jun 10th, 2021. No vaccine percentages are available from Jun 1st, 2021, so the earliest vaccine coverage was chosen from the database. No vaccine percentages are available from Feb 13th, 2023, so the latest vaccine coverage was chosen from the database. The maximum of the percentage from this calculation and the calculation from **Vaccine coverage: Full course** was used.<br><br>***Methodology***<br>Set country as “China”. Set Metric to “People vaccinated”. Select “Relative to Population”.<br><br>Jun 10th, 2021: 43.62%<br>Feb 9th, 2023: 91.89%<br><br>Vaccine coverage: 91.89% - 43.62% = 48.27% |
| Vaccine coverage: Full course<br><br>ChinaWHO!B3:H3 | https://ourworldindata.org/covid-vaccinations<br><br>The coverage for individuals who took the full course of the vaccine during the 20.5-month period of vaccine durability was calculated by subtracting the cumulative percentage from Feb 9th, 2023, by the cumulative percentage from Aug 12th, 2021. No vaccine percentages are available from Jun 1st, 2021, so the earliest vaccine coverage was chosen from the database. No vaccine percentages are available from Feb 13th, 2023, so the latest vaccine coverage was chosen from the database.<br><br>***Methodology***<br>Set country as “China”. Set Metric to “People fully vaccinated”. Select “Relative to Population”. |

| | |
|---|---|
| | Aug 12th, 2021: 54.50%<br>Feb 9th, 2023: 89.54%<br><br>Vaccine coverage: 89.54% - 54.40% = 35.04% |
| Infection-derived immunity<br><br>ChinaWHO!B4:H4 | https://www.nature.com/articles/d41586-023-01872-7<br><br>Although infection-derived immunity was calculated using cumulative cases in all other countries, because of the confusion on the cumulative number of cases in China an alternate source was used. The article writes that approximately 85% of China's population was infected in December 2022, which is corroborated by other news outlets. |
| Prevalence<br><br>ChinaWHO!B5:H5 | Prevalence was calculated by dividing the weekly cases by the total population.<br><br>Prevalence = 94,489/1,409,778,724 = 0.00006702 |
| Population size<br><br>ChinaWHO!B6:H6 | http://www.stats.gov.cn/sj/ndsj/2021/indexeh.htm<br><br>The population sizes for each age group was extracted from the China National Bureau of Statistics through the China Statistical Yearbook 2021 (2-17: Population by Age and Gender (2020)).<br><br>***Methodology***<br>*Notes*<br>The age group of 15-19 was separated across the <18 and 18-29 groups by assuming an equal number of individuals for each age.<br><br><18: 77,883,888 + 90,244,056 + 85,255,994 + 72,684,140*3/5 = 296994422<br>18-29: 72,684,140*2/5 + 74,941,675 + 91,847,332 = 195862663<br>30-39: 124,145,190 + 99012932 = 223,158,122<br>40-49: 92,955,330 + 114224887 = 207,180,217<br>50-59: 121,164,296 + 101,400,786 = 222,565,082<br>60+: 73,382,938 + 74,005,560 + 49,590,036 + 31,238,849 + 35,800,835 = 264,018,218<br><br>Total population: 1,409,778,724 |
| Cumulative Cases<br><br>ChinaWHO!B8 | https://covid19.who.int/region/wpro/country/cn<br><br>The cumulative cases during the 21.5-month period of infection-derived immunity durability was calculated by subtracting the cumulative number of cases from Feb 19th, 2023, by the |

| | |
|---|---|
| | cumulative number of cases from May 1st, 2021.<br><br>***Methodology***<br>Select "Cumulative". Select "Daily". Hover over the desired date and choose the number for confirmed cases.<br><br>May 1st, 2021: 103,649<br>Feb 19th, 2023: 98,904,475<br><br>Cumulative cases: 98,904,475 - 103,649 = 98,800,826 |
| Weekly Cases<br><br>ChinaWHO!B9 | https://covid19.who.int/region/wpro/country/cn<br><br>The weekly cases was extracted from the official WHO database for the week of Feb 13th to Feb 19th.<br><br>***Methodology***<br>Select "Daily Change". Select "Weekly". Hover over Feb 13th, 2023, and choose the number for confirmed cases.<br><br>Weekly cases: 94,489 |
| China Inbounding Travellers (2019)<br><br>ChinaWHO!B10 | https://www.wta-web.org/wp-content/uploads/2022/03/China-Inbound-Tourism-Development-Report.pdf<br><br>The World Tourism Alliance estimated the inbound travelers to China was approximately 145,307,800 in 2019 (page 13). |
| China Estimated Inbounding Travellers (Feb 2023)<br><br>ChinaWHO!B11 | https://www.mckinsey.com/industries/travel-logistics-and-infrastructure/our-insights/what-to-expect-from-chinas-travel-rebound#/<br><br>Because of the lack of information on inbounding travelers to China in February 2023, an estimate from McKinsey and Company was employed, predicting that China's tourism recovery would follow a trajectory similar to that of Hong Kong. Using the graph, roughly two million travelers will visit China in February 2023. |

**China (Surveillance Data)**

| Data | Link and Extraction Methods |
|---|---|
| Prevalence<br><br>ChinaSurveillance!B5:H5 | https://www.ncbi.nlm.nih.gov/pmc/articles/PMC10184382/<br><br>In a study by Fu *et al.*, out of an online survey filled out buy 2,391 Chinese participants, an estimated 0.1% self-reported as being actively infectious between February 2nd to February 4th. |

**Japan**

| Data | Link and Extraction Methods |
|---|---|
| Vaccine coverage: At least one dose<br><br>Japan!B2:H2 | https://ourworldindata.org/covid-vaccinations<br><br>The coverage for individuals who took at least one dose of the vaccine during the 20.5-month period of vaccine durability was calculated by subtracting the cumulative percentage from Feb 13th, 2023, by the cumulative percentage from Jun 1st, 2021. The maximum of the percentage from this calculation and the calculation from **Vaccine coverage: Full course** was used.<br><br>***Methodology***<br>Set country as "Japan". Set Metric to "People vaccinated". Select "Relative to Population".<br><br>Jun 1st, 2021: 10.87%<br>Feb 13th, 2023: 84.43%<br><br>Vaccine coverage: 84.43% - 10.87% = 73.56% |
| Vaccine coverage: Full course<br><br>Japan!B3:H3 | https://ourworldindata.org/covid-vaccinations<br><br>The coverage for individuals who took the full course of the vaccine during the 20.5-month period of vaccine durability was calculated by subtracting the cumulative percentage from Feb 13th, 2023, by the cumulative percentage from Jun 1st, 2021.<br><br>***Methodology***<br>Set country as "Japan". Set Metric to "People fully vaccinated". Select "Relative to Population".<br><br>Jun 1st, 2021: 3.47%<br>Feb 13th, 2023: 83.34%<br><br>Vaccine coverage: 83.34% - 3.47% = 79.87% |
| Infection-derived immunity<br><br>Japan!B4:H4 | Infection-derived immunity was calculated by dividing the number of cumulative cases over the last 21.5 months, over the duration of the durability of immunity, by the total population.<br><br>Infection-derived immunity = 32,479,468/124,862,000 = 0.2601229 |
| Prevalence | Prevalence was calculated by dividing the weekly cases by the total population. |

| | |
|---|---|
| Japan!B5:H5 | Prevalence = 141,082/124,862,000 = 0.0011300 |
| Population size<br><br>Japan!B6:H6 | https://www.stat.go.jp/english/data/jinsui/tsuki/index.html<br><br>The population sizes for each age group were extracted from the Statistics Bureau of Japan Monthly Report.<br><br>***Methodology***<br>*Notes*<br>The age group of 15-19 was separated across the <18 and 18-29 groups by assuming an equal number of individuals for each age.<br><br><18: 4,223,000 + 4,929,000 + 5297000 + 5,516,000*3/5 = 17,758,600<br>18-29: 5,516,000*2/5 + 6,266,000 + 6,420,000 = 14,892,400<br>30-39: 6,434,000 + 7,189,000 = 13,623,000<br>40-49: 7,915,000 + 9,415,000 = 17,330,000<br>50-59: 9,474,000 + 8,112,000 = 17,586,000<br>60+: 7,462,000 + 7,484,000 + 9,245,000 + 7,111,000 + 5,738,000 + 3,969,000 + 2,008,000 + 567,000 + 88,000 = 43,672,000<br><br>Total population: 124,862,000 |
| Inbounding Travellers (China to Japan) (Feb 2023)<br><br>Japan!B8, Japan!B9 | https://www.tourism.jp/en/tourism-database/stats/inbound/<br><br>Japan’s Tourism department reports the number of tourists from China who visited Japan monthly. 36,200 visitors came in Feb 2023 (1293 per day) |
| Outbounding Travellers (Japan to China) (2018)<br><br>Japan!B10 | http://www.stats.gov.cn/sj/ndsj/2019/indexeh.htm<br><br>The number of travelers from Japan to China was extracted from the China National Bureau of Statistics through the China Statistical Yearbook 2019 (17-13: Number of Oversea Visitor Arrivals by Country/Region).<br><br>Outbound Travelers: 2,691,400 |
| Outbounding Travellers (Japan to China) (2023)<br><br>Japan!B11, Japan!B12 | Using the McKinsey estimate, we can scale the estimated number of outbound travelers from Japan to China.<br><br>Feb 2023 = 2,691,400*2,000,000/145,307,800 = 37044<br>Daily = 37044/28 = 1323 |
| Length of Stay (China to Japan) (2021) | https://statistics.jnto.go.jp/en/graph/#graph--average--length--of--stay |

| | |
|---|---|
| Japan!B13 | Data was derived from Japan's official Tourism Department. Select "China" for Country/Area.<br><br>Length of Stay: 47.4 days |
| Length of Stay (Japan to China) (2019)<br><br>Japan!B154 | https://www.wta-web.org/wp-content/uploads/2022/03/China-Inbound-Tourism-Development-Report.pdf<br><br>The World Tourism Alliance estimated the average length of stay for inbound tourists to China was around 9.2 days (page 49). |
| Cumulative Cases<br><br>Japan!B15 | https://covid19.who.int/region/wpro/country/jp<br><br>The number of cumulative cases during the 21.5-month period of infection-derived immunity durability was calculated by subtracting the cumulative number of cases from Feb 19th, 2023, by the cumulative number of cases from May 1st, 2021.<br><br>***Methodology***<br>Select "Cumulative". Select "Daily". Hover over the desired data and choose the number for confirmed cases.<br><br>May 1st, 2021: 597,225<br>Feb 19th, 2023: 33,076,693<br><br>Cumulative Cases: 33,076,693 - 597,225 = 32,479,468 |
| Weekly Cases<br><br>Japan!B16, Japan!B17 | https://covid19.who.int/region/wpro/country/jp<br><br>The weekly cases was extracted from the official WHO database for the week of Feb 13th to Feb 19th.<br><br>***Methodology***<br>Select "Daily Change". Select "Weekly". Hover over Feb 13th, 2023, and choose the number for confirmed cases.<br><br>Weekly cases: 141,082<br>Daily Incidence = 141,082/7 = 20,155 |

**South Korea**

| Data | Link and Extraction Methods |
|---|---|
| Vaccine coverage: At least one dose | https://ourworldindata.org/covid-vaccinations |

| | |
|---|---|
| SouthKorea!B2:H2 | The coverage for individuals who took at least one dose of the vaccine during the 20.5-month period of vaccine durability was calculated by subtracting the cumulative percentage from Feb 13th, 2023, by the cumulative percentage from Jun 1st, 2021. The maximum of the percentage from this calculation and the calculation from **Vaccine coverage: Full course** was used.<br><br>***Methodology***<br>Set country as "South Korea". Set Metric to "People vaccinated". Select "Relative to Population".<br><br>Jun 1st, 2021: 12.14%<br>Feb 13th, 2023: 86.41%<br><br>Vaccine coverage: 86.41% - 12.14% = 74.27% |
| Vaccine coverage: Full course<br><br>SouthKorea!B3:H3 | https://ourworldindata.org/covid-vaccinations<br><br>The coverage for individuals who took the full course of the vaccine during the 20.5-month period of vaccine durability was calculated by subtracting the cumulative percentage from Feb 13th, 2023, by the cumulative percentage from Jun 1st, 2021.<br><br>***Methodology***<br>Set country as "South Korea". Set Metric to "People fully vaccinated". Select "Relative to Population".<br><br>Jun 1st, 2021: 4.27%<br>Feb 13th, 2023: 85.61%<br><br>Vaccine coverage: 85.61% - 4.27% = 81.34% |
| Infection-derived immunity<br><br>SouthKorea!B4:H4 | Infection-derived immunity was calculated by dividing the number of cumulative cases over the last 21.5 months, over the duration of the durability of immunity, by the total population.<br><br>Infection-derived immunity = 30,306,339/51,815,797 = 0.5848861 |
| Prevalence<br><br>SouthKorea!B5:H5 | Prevalence was calculated by dividing the weekly cases by the total population.<br><br>Prevalence = 79,372/51,815,797 = 0.0015318 |
| Population size<br><br>SouthKorea!B6:H6 | https://www.populationpyramid.net/republic-of-korea/2022/<br><br>The population sizes for each age group was extracted from |

| | |
|---|---|
| | PopulationPyramid.net, which uses information from the United Nations.<br><br>***Methodology***<br>*Notes*<br>The age group of 15-19 was separated across the <18 and 18-29 groups by assuming an equal number of individuals for each age.<br><br><18: 1,553,522 + 2,137,592 + 2,304,825 + 2,300,682*3/5 = 7,376,348<br>18-29: 2,300,682*2/5 + 3,083,300 + 3624178 = 7,627,751<br>30-39: 3,405,340 + 3,518,709 = 6,924,049<br>40-49: 4,033,819 + 4,057,848 = 8,091,667<br>50-59: 4,489,610 + 4,091,813 = 8,581,423<br>60+: 4,151,390 + 3,092,536 + 2,152,607 + 1,592,179 + 1,238,961 + 657,693 + 260,012 + 60,335 + 8,846 = 13,214,559<br><br>Total population: 51,815,797 |
| Inbounding Travellers (China to South Korea) (Dec 2022)<br><br>SouthKorea!B8, SouthKorea!B9 | https://www.koreatimes.co.kr/www/culture/2023/04/141_348163.html<br><br>The Korea Times reports that according to the official Korean Tourism Organization, 45,900 Chinese travelers visited in Feb 2023 (1,639 per day). |
| Outbounding Travellers (South Korea to China) (2018)<br><br>SouthKorea!B10 | http://www.stats.gov.cn/sj/ndsj/2019/indexeh.htm<br><br>The number of travelers from South Korea to China was extracted from the China National Bureau of Statistics through the China Statistical Yearbook 2019 (17-13: Number of Oversea Visitor Arrivals by Country/Region).<br><br>Outbound Travelers: 4,193,500 |
| Outbounding Travellers (South Korea to China) (2023)<br><br>SouthKorea!B11, SouthKorea!B12 | Using the McKinsey estimate, we can scale the estimated number of outbound travelers from South Korea to China.<br><br>Feb 2023 = 4,193,500*2,000,000/145,307,800 = 57,719<br>Daily = 57,719/28 = 2061.4 |
| Length of Stay (China to South Korea) (2021)<br><br>SouthKorea!B13 | https://www.statista.com/statistics/1133384/south-korea-average-length-of-stay-for-visitors-by-origin/<br><br>Chinese travelers stay in South Korea for an estimated 41.9 days. |

| | |
|---|---|
| Length of Stay (South Korea to China) (2019)<br><br>SouthKorea!B14 | https://www.wta-web.org/wp-content/uploads/2022/03/China-Inbound-Tourism-Development-Report.pdf<br><br>The World Tourism Alliance estimated the average length of stay for inbound tourists to China was around 9.2 days (page 49). |
| Cumulative Cases<br><br>SouthKorea!B15 | https://covid19.who.int/region/wpro/country/kr<br><br>The cumulative cases during the 21.5-month period of infection-derived immunity durability was calculated by subtracting the cumulative number of cases from Feb 19th, 2023, by the cumulative number of cases from May 1st, 2021.<br><br>***Methodology***<br>Select “Cumulative”. Select “Daily”. Hover over the desired data and choose the number of confirmed cases.<br><br>May 1st, 2021: 123,232<br>Feb 19th, 2023: 30,429,571<br><br>Cumulative Cases: 30,429,571 - 123,232 = 30,306,339 |
| Weekly Cases<br><br>SouthKorea!B16, SouthKorea!B17 | https://covid19.who.int/region/wpro/country/kr<br><br>The number of weekly cases was extracted from the official WHO database for the week of Feb 13th to Feb 19th.<br><br>***Methodology***<br>Select “Daily Change”. Select “Weekly”. Hover over Feb 13th, 2023, and choose the number for confirmed cases.<br><br>Weekly Cases: 79,372<br>Daily Incidence: 79,372/7 = 11,339 |

**Singapore**

| Data | Link and Extraction Methods |
|---|---|
| Vaccine coverage: At least one dose<br><br>Singapore!B2:H2 | https://ourworldindata.org/covid-vaccinations<br><br>The coverage for individuals who took at least one dose of the vaccine during the 20.5-month period of vaccine durability was calculated by subtracting the cumulative percentage from Jan 30th, 2023, by the cumulative percentage from Jun 1st, 2021. No vaccine percentages are available from Feb 13th, 2023, so the latest vaccine coverage was chosen from the database. The maximum of the |

| | |
|---|---|
| | percentage from this calculation and the calculation from **Vaccine coverage: Full course** was used.<br><br>***Methodology***<br>Set country as “Singapore”. Set Metric to “People vaccinated”. Select “Relative to Population”.<br><br>Jun 1st, 2021: 40.44%<br>Jan 30th, 2023: 91.55%<br><br>Vaccine coverage: 91.55% - 40.44% = 51.11% |
| Vaccine coverage: Full course<br><br>Singapore!B3:H3 | https://ourworldindata.org/covid-vaccinations<br><br>The coverage for individuals who took the full course of the vaccine during the 20.5-month period of vaccine durability was calculated by subtracting the cumulative percentage from Jan 30th, 2023, by the cumulative percentage from Jun 1st, 2021. No vaccine percentages are available from Feb 13th, 2023, so the latest vaccine coverage was chosen from the database.<br><br>***Methodology***<br>Set country as “Singapore”. Set Metric to “People fully vaccinated”. Select “Relative to Population”.<br><br>Jun 1st, 2021: 31.26%<br>Jan 30th, 2023: 90.84%<br><br>Vaccine coverage: 90.84% - 31.26% = 59.58% |
| Infection-derived immunity<br><br>Singapore!B4:H4 | Infection-derived immunity was calculated by dividing the number of cumulative cases over the last 21.5 months, over the duration of the durability of immunity, by the total population.<br><br>Infection-derived immunity = 2,160,827/4,073,239 = 0.5304935 |
| Prevalence<br><br>Singapore!B5:H5 | Prevalence was calculated by dividing the weekly cases by the total population.<br><br>Prevalence = 3,849/4,073,239 = 0.0009449 |
| Population size<br><br>Singapore!B6:H6 | https://tablebuilder.singstat.gov.sg/table/TS/M810011<br><br>The population sizes for each age group were extracted from the official Singapore census data. |

| | |
|---|---|
| | ***Methodology***<br>*Notes*<br>The age group of 15-19 was separated across the <18 and 18-29 groups by assuming an equal number of individuals for each age.<br><br><18: 178,085 + 201,360 + 202,379 + 206,749*3/5 = 705,873<br>18-29: 206,749*2/5 + 233,303 + 280,082 = 596,085<br>30-39: 317,153 + 290,981 = 608,134<br>40-49: 299,871 + 304,317 = 604,188<br>50-59: 292,984 + 299,835 = 592,819<br>60+: 288,007 + 678,133 = 966,140<br><br>Total population: 4,073,239 |
| Inbounding Travellers (China to Singapore) (Feb 2023)<br><br>Singapore!B8, Singapore!B9 | [https://www.singstat.gov.sg/publications/reference/ebook/industry/tourism](https://www.singstat.gov.sg/publications/reference/ebook/industry/tourism)<br><br>Singapore's Tourism department reports the number of tourists from China who visited Singapore monthly. 35,312 visitors from mainland China came in Feb 2023 (1,261 per day) |
| Outbounding Travellers (Singapore to China) (2018)<br><br>Singapore!B10 | [http://www.stats.gov.cn/sj/ndsj/2019/indexeh.htm](http://www.stats.gov.cn/sj/ndsj/2019/indexeh.htm)<br><br>The number of travelers from Singapore to China was extracted from the China National Bureau of Statistics through the China Statistical Yearbook 2019 (17-13: Number of Oversea Visitor Arrivals by Country/Region).<br><br>Outbound Travelers: 978,400 |
| Outbounding Travellers (Singapore to China) (2023)<br><br>Singapore!B11, Singapore!B12 | Using the McKinsey estimate, we can scale the estimated number of outbound travelers from Singapore to China.<br><br>Feb 2023 = 978,400*2,000,000/145,307,800 = 13,467<br>Daily = 13,467/28 = 481 |
| Length of Stay (China to Singapore) (2022)<br><br>Singapore!B13 | [https://www.scmp.com/news/asia/southeast-asia/article/3207085/singapore-expects-billions-more-tourism-dollars-china-boost](https://www.scmp.com/news/asia/southeast-asia/article/3207085/singapore-expects-billions-more-tourism-dollars-china-boost)<br><br>Executives from Singapore's Tourism Board report that the average Chinese traveler stayed in Singapore for 4.81 days. |
| Length of Stay (Singapore to China) (2019) | [https://www.wta-web.org/wp-content/uploads/2022/03/China-Inbound-Tourism-Development-Report.pdf](https://www.wta-web.org/wp-content/uploads/2022/03/China-Inbound-Tourism-Development-Report.pdf)<br><br>The World Tourism Alliance estimated the average length of stay for |

| | |
|---|---|
| Singapore!B14 | inbound tourists to China was around 9.2 days (page 49). |
| Cumulative Cases<br><br>Singapore!B15 | https://covid19.who.int/region/wpro/country/sg<br><br>The cumulative cases during the 21.5-month period of infection-derived immunity durability was calculated by subtracting the cumulative number of cases from Feb 19th, 2023, by the cumulative number of cases from May 1st, 2021.<br><br>***Methodology***<br>Select "Cumulative". Select "Daily". Hover over the desired data and choose the number for confirmed cases.<br><br>May 1st, 2021: 61,179<br>Feb 19th, 2023: 2,222,006<br><br>Cumulative Cases: 2,222,006 - 61,179 = 2,160,827 |
| Weekly Cases<br><br>Singapore!B16, Singapore!B17 | https://covid19.who.int/region/wpro/country/sg<br><br>The weekly cases was extracted from the official WHO database for the week of Feb 20th to Feb 27th. This week was used instead of the week of Feb 13th to 19th because the latter was very significantly lower than the other weekly case numbers, and potentially an outlier.<br><br>***Methodology***<br>Select "Daily Change". Select "Weekly". Hover over Feb 20th, 2023, and choose the number for confirmed cases.<br><br>Weekly cases: 3,849<br>Daily Incidence: 3,849/7 = 550 |

**England**

| Data | Link and Extraction Methods |
|---|---|
| Vaccine coverage: At least one dose<br><br>England!B2:H2 | https://ourworldindata.org/covid-vaccinations<br><br>The coverage for individuals who took at least one dose of the vaccine during the 20.5-month period of vaccine durability was calculated by subtracting the cumulative percentage from Feb 13th, 2023, by the cumulative percentage from Jun 1st, 2021. The maximum of the percentage from this calculation and the calculation from **Vaccine coverage: Full course** was used. |

| | |
|---|---|
| | ***Methodology***<br>Set country as “England”. Set Metric to “People vaccinated”. Select “Relative to Population”.<br><br>Jun 1st, 2021: 58.51%<br>Feb 13th, 2023: 80.29%<br><br>Vaccine coverage: 80.29% - 58.21% = 22.08% |
| Vaccine coverage: Full course<br><br>England!B3:H3 | [https://ourworldindata.org/covid-vaccinations](https://ourworldindata.org/covid-vaccinations)<br><br>The coverage for individuals who took the full course of the vaccine during the 20.5-month period of vaccine durability was calculated by subtracting the cumulative percentage from Feb 13th, 2023, by the cumulative percentage from Jun 1st, 2021.<br><br>***Methodology***<br>Set country as “England”. Set Metric to “People fully vaccinated”. Select “Relative to Population”.<br><br>Jun 1st, 2021: 39.17%<br>Feb 13th, 2023: 75.92%<br><br>Vaccine coverage: 75.92% - 39.17% = 36.75% |
| Infection-derived immunity<br><br>England!B4:H4 | Infection-derived immunity was calculated by dividing the number of cumulative cases over the last 21.5 months, over the duration of the durability of immunity, by the total population.<br><br>Infection-derived immunity: 16,650,385/56,536,419 = 0.2945072 |
| Prevalence<br><br>England!B5:H5 | Prevalence was calculated by dividing the weekly cases by the total population.<br><br>Prevalence: 22,380/56,536,419 = 0.0003959 |
| Population size<br><br>England!B6:H6 | [https://www.ons.gov.uk/peoplepopulationandcommunity/populationandmigration/populationestimates/bulletins/populationandhouseholdestimatesenglandandwales/census2021](https://www.ons.gov.uk/peoplepopulationandcommunity/populationandmigration/populationestimates/bulletins/populationandhouseholdestimatesenglandandwales/census2021)<br><br>The population sizes for each age group was extracted from official England census data.<br><br>***Methodology***<br><br><18: 579,315 + 601,274 + 614,109 + 623,873 + 639,646 + 658,513 |

| | |
|---|---|
| | + 653,208 + 658,120 + 675,503 + 694,918 + 696,488 + 688,076 + 683,067 + 689,261 + 663,078 + 649,499 + 648,066 + 645,642 = 11,761,656<br>18-29: 637,270 + 641,579 + 650,705 + 661,796 + 684,929 + 695,509 + 714,474 + 705,555 + 720,077 + 744,015 + 749,401 + 775,860 = 8,381,170<br>30-39: 789,926 + 791,601 + 788,320 + 799,021 + 781,498 + 776,514 + 774,632 + 750,999 + 751,814 + 748,949 = 7,753,274<br>40-49: 757,542 + 760,845 + 729,833 + 679,655 + 667,273 + 681,401 + 695,685 + 704,152 + 731,536 + 759,044 = 7,166,966<br>50-59: 786,831 + 766,990 + 783,664 + 781,179 + 785,653 + 781,563 + 782,568 + 770,902 + 751,858 + 733,101 = 7,724,309<br>60+: 704,980 + 674,965 + 655,837 + 636,312 + 608,797 + 583,874 + 559,535 + 558,458 + 546,304 + 528,480 + 528,807 + 533,882 + 543,415 + 570,305 + 613,713 + 464,281 + 444,154 + 435,404 + 395,427 + 345,640 + 301,313 + 305,184 + 293,030 + 274,127 + 249,035 + 223,864 + 200,094 + 173,198 + 152,119 + 135,074 + 509,436 = 13,749,044<br><br>Total population: 56,536,419 |
| Inbounding Travellers (China to England) (Q4 2022)<br><br>England!B8, England!B9 | https://www.oxfordeconomics.com/resource/china-travel-recovery-timings-are-clear-but-magnitude-remains-uncertain-for-2023/<br><br>https://www.visitbritain.org/markets/china<br><br>In 2018, 686,433 travelers from China visited England. This was calculated by subtracting the total number of visitors to the UK to the visitors to other locations (883,073 - 171,650 - 17,440 - 1,310 - 6,240 = 686,433).<br><br>The recovery rate of Chinese travelers is expected to be about 48%, or that 686,433*0.48 = 329,488 travelers will come from China to England in 2023 (903 per day). |
| Outbounding Travellers (England to China) (2019)<br><br>England!B10 | https://www.visitscotland.org/research-insights/about-our-visitors/international/china<br><br>An estimated 598,000 individuals from England traveled to China in 2019. The number of travelers from England to China was estimated by subtracting the number of travelers from the UK by the number of travelers from Scotland (646,000 - 48,000 = 598,000). |
| Outbounding Travellers (England to China) (2023) | Using the McKinsey estimate, we can scale the estimated number of outbound travelers from England to China. |

| | |
|---|---|
| England!B11,<br>England!B12 | Feb 2023: 598,000*2,000,000/145,307,800 = 8,231<br>Daily: 8,231/28 = 294 |
| Length of Stay (China to England) (2018)<br><br>England!B13 | https://www.visitbritain.org/markets/china<br><br>The average length of stay of Chinese travelers in England can be estimated using the average length of stay of Chinese travelers in the UK. The average length of stay is around 16.33 days. |
| Length of Stay (England to China) (2019)<br><br>England!B14 | https://www.visitscotland.org/research-insights/about-our-visitors/international/china<br><br>The average length of stay of English travelers in China can be estimated using the average length of stay of all UK travelers in China. The average length of stay is around 24.6 days. |
| Cumulative Cases<br><br>England!B15 | https://coronavirus.data.gov.uk/details/cases?areaType=nation&areaName=England<br><br>The cumulative cases during the 21.5-month period of infection-derived immunity durability was calculated by subtracting the cumulative number of cases from Feb 19th, 2023, by the cumulative number of cases from May 1st, 2021.<br><br>***Methodology***<br>On the graph named "Cases by specimen date", click the "Total" tab. Hover over the desired date and choose the number of cases.<br><br>May 1st, 2021: 3,948,017<br>Feb 19th, 2023: 20,598,402<br><br>Cumulative Cases: 20,598,402 - 3,948,017 = 16,650,385 |
| Weekly Cases<br><br>England!B16,<br>England!B17 | https://coronavirus.data.gov.uk/details/cases?areaType=nation&areaName=England<br><br>The weekly cases was extracted from the official UK COVID database for the week of Feb 13th to Feb 19th.<br><br>***Methodology***<br>On the graph named "Cases by specimen date", click the "Total" tab. Hover over the desired date and choose the number of cases.<br><br>Feb 13th, 2023: 20,576,022<br>Feb 19th, 2023: 20,598,402 |

| | |
|---|---|
| | Weekly Cases: 20,598,402 - 20,576,022 = 22,380<br>Daily Incidence: 22,380/7 = 3,197 |

**Scotland**

| Data | Link and Extraction Methods |
|---|---|
| Vaccine coverage: At least one dose<br><br>Scotland!B2:H2 | https://ourworldindata.org/covid-vaccinations<br><br>The coverage for individuals who took at least one dose of the vaccine during the 20.5-month period of vaccine durability was calculated by subtracting the cumulative percentage from Feb 13th, 2023, by the cumulative percentage from Jun 1st, 2021. No vaccine percentages are available from Feb 13th, 2023, so the latest vaccine coverage was chosen from the database. The maximum of the percentage from this calculation and the calculation from **Vaccine coverage: Full course** was used.<br><br>***Methodology***<br>Set country as "Scotland". Set Metric to "People vaccinated". Select "Relative to Population".<br><br>Jun 1st, 2021: 60.12%<br>Sep 11th, 2022: 83.23%<br><br>Vaccine coverage: 83.23% - 60.12% = 23.11% |
| Vaccine coverage: Full course<br><br>Scotland!B3:H3 | https://ourworldindata.org/covid-vaccinations<br><br>The coverage for individuals who took the full course of the vaccine during the 20.5-month period of vaccine durability was calculated by subtracting the cumulative percentage from Feb 13th, 2023, by the cumulative percentage from Jun 1st, 2021. No vaccine percentages are available from Feb 13th, 2023, so the latest vaccine coverage was chosen from the database.<br><br>***Methodology***<br>Set country as "Scotland". Set Metric to "People fully vaccinated". Select "Relative to Population".<br><br>Jun 1st, 2021: 38.53%<br>Sep 11th, 2022: 78.41%<br><br>Vaccine coverage: 78.41% - 38.53% = 39.88% |

| | |
|---|---|
| Infection-derived immunity<br><br>Scotland!B4:H4 | Infection-derived immunity was calculated by dividing the number of cumulative cases over the last 21.5 months, over the duration of the durability of immunity, by the total population.<br><br>Infection-derived immunity: 1,927,983/5,479,900 = 0.3518281 |
| Prevalence<br><br>Scotland!B5:H5 | Prevalence was calculated by dividing the weekly cases by the total population.<br><br>Prevalence: 1,655/5,479,900 = 0.0003020 |
| Population size<br><br>Scotland!B6:H6 | https://www.ons.gov.uk/peoplepopulationandcommunity/populationandmigration/populationestimates/datasets/populationestimatesforukenglandandwalesscotlandandnorthernireland<br><br>The population sizes for each age group was extracted from official Scotland census data.<br><br>***Methodology***<br><br><18: 46,782 + 49,017 + 51,478 + 53,317 + 54,843 + 57,070 + 57,945 + 58,262 + 59,490 + 60,960 + 62,868 + 59,950 + 61,557 + 61,334 + 58,857 + 57,792 + 57,280 + 56,179 = 1,024,981<br>18-29: 55,074 + 57,305 + 61,314 + 63,526 + 66,277 + 69,034 + 71,827 + 71,242 + 71,264 + 73,491 + 75,198 + 79,523 = 815,075<br>30-39: 79,853 + 76,958 + 76,395 + 76,227 + 73,900 + 73,404 + 72,855 + 70,423 + 71,179 + 72,113 = 743,307<br>40-49: 71,460 + 69,923 + 67,428 + 62,504 + 61,236 + 64,443 + 64,670 + 65,770 + 69,572 + 73,843 = 670,849<br>50-59: 76,442 + 75,420 + 78,255 + 79,663 + 80,011 + 79,389 + 81,987 + 80,973 + 80,622 + 78,439 = 791,201<br>60+: 76,225 + 73,417 + 72,636 + 70,250 + 68,098 + 65,670 + 62,371 + 61,188 + 59,360 + 56,835 + 56,692 + 56,433 + 57,056 + 58,119 + 61,854 + 45,663 + 42,249 + 42,149 + 39,250 + 34,988 + 31,336 + 30,894 + 29,041 + 27,044 + 24,360 + 21,934 + 19,692 + 16,983 + 14,476 + 12,903 + 45,321 = 1,434,487<br><br>Total population: 5,479,900 |
| Inbounding Travellers (China to Scotland) (2019)<br><br>Scotland!B8, Scotland!B9 | https://www.oxfordeconomics.com/resource/china-travel-recovery-timings-are-clear-but-magnitude-remains-uncertain-for-2023/<br><br>https://www.visitbritain.org/markets/china<br><br>In 2018, 171,650 travelers from China visited Scotland. The recovery rate of Chinese travelers is expected to be about 48%, |

| | |
|---|---|
| | meaning 171,650*0.48 = 82,392 travelers will come from China to Scotland in 2023 (226 per day). |
| Outbounding Travellers (Scotland to China) (2019)<br><br>Scotland!B10 | https://www.visitscotland.org/research-insights/about-our-visitors/international/china<br><br>In 2019, an approximate 48,000 people from Scotland visited China. |
| Outbounding Travellers (Scotland to China) (2023)<br><br>Scotland!B11, Scotland!B12 | Using the McKinsey estimate, we can scale the estimated number of outbound travelers from Scotland to China.<br><br>Feb 2023: 48,000*2,000,000/145,307,800 = 661<br>Daily: 661/28 = 23.6 |
| Length of Stay (China to Scotland) (2019)<br><br>Scotland!B13 | https://www.visitscotland.org/research-insights/about-our-visitors/international/china<br><br>In 2019, the average stay of Chinese visitors in Scotland was 11.9 days. |
| Length of Stay (Scotland to China) (2019)<br><br>Scotland!B14 | https://www.visitscotland.org/research-insights/about-our-visitors/international/china<br><br>The average length of stay of Scottish travelers in China can be estimated using the average length of stay of all UK travelers in China. The average length of stay is around 24.6 days. |
| Cumulative Cases<br><br>Scotland!B15 | https://scotland.shinyapps.io/phs-respiratory-covid-19/<br><br>The number of cumulative cases during the 21.5-month period of infection-derived immunity durability was calculated from Feb 19th, 2023 to May 1st, 2021.<br><br>***Methodology***<br>The data for the Reported COVID-19 Cases was downloaded, then summed from May 1st, 2021 to Feb 19th, 2023.<br><br>Cumulative Cases: 1,927,983 |
| Weekly Cases<br><br>Scotland!B16, Scotland!B17 | https://scotland.shinyapps.io/phs-respiratory-covid-19/<br><br>The number of weekly cases was extracted from the official Scotland COVID database for the week of Feb 13th to Feb 19th.<br><br>***Methodology*** |

| | |
|---|---|
| | The data for the Reported COVID-19 Cases was downloaded, then summed from Feb 13th, 2023 to Feb 19th, 2023.<br><br>Weekly Cases: 1,655<br>Daily Incidence: 1,655/7 = 236 |

**France**

| Data | Link and Extraction Methods |
|---|---|
| Vaccine coverage: At least one dose<br><br>France!B2:H2 | https://ourworldindata.org/covid-vaccinations<br><br>The coverage for individuals who took at least one dose of the vaccine during the 20.5-month period of vaccine durability was calculated by subtracting the cumulative percentage from Feb 13th, 2023, by the cumulative percentage from Jun 1st, 2021. The maximum of the percentage from this calculation and the calculation from **Vaccine coverage: Full course** was used.<br><br>***Methodology***<br>Set the country as “France”. Set Metric to “People vaccinated”. Select “Relative to Population”. Select “Cumulative” for the interval.<br><br>Jun 1st, 2021: 39.59%<br>Feb 13th, 2023: 80.61%<br><br>Vaccine coverage: 80.61% - 39.59% = 41.02% |
| Vaccine coverage: Full course<br><br>France!B3:H3 | https://ourworldindata.org/covid-vaccinations<br><br>The coverage for individuals who took the full course of the vaccine during the 20.5-month period of vaccine durability was calculated by subtracting the cumulative percentage from Feb 13th, 2023, by the cumulative percentage from Jun 1st, 2021.<br><br>***Methodology***<br>Set the country as “France”. Set Metric to “People fully vaccinated”. Select “Relative to Population”.<br><br>Jun 1st, 2021: 17.18%<br>Feb 13th, 2023: 78.41%<br><br>Vaccine coverage: 78.41% - 17.18% = 61.23% |
| Infection-derived | Infection-derived immunity was calculated by dividing the number |

| | |
|---|---|
| immunity<br><br>France!B4:H4 | of cumulative cases over the last 21.5 months, over the duration of the durability of immunity, by the total population.<br><br>Infection-derived immunity: 33,277,148/67,063,703 = 0.4962021 |
| Prevalence<br><br>France!B5:H5 | Prevalence was calculated by dividing the weekly cases by the total population.<br><br>Prevalence: 23,862/67,063,703 = 0.0003558 |
| Population size<br><br>France!B6:H6 | https://www.insee.fr/en/statistiques/2382597?sommaire=2382613<br><br>***Methodology***<br>*Notes*<br><br><18: 706,382 + 716,159 + 729,139 + 749,142 + 770,897 + 795,049 + 801,336 + 818,973 + 824,266 + 844,412 + 836,610 + 841,774 + 833,484 + 847,250 + 828,874 + 828,224 + 825,535 + 824,243 = 14,421,749<br>18-29: 830,859 + 832,135 + 778,595 + 767,419 + 738,255 + 741,493 + 731,720 + 709,814 + 710,229 + 747,365 + 762,740 + 783,278 = 9,133,902<br>30-39: 793,756 + 805,709 + 809,462 + 824,388 + 823,154 + 817,616 + 809,113 + 860,183 + 868,514 + 876,362 = 8,288,257<br>40-49: 830,619 + 812,560 + 815,529 + 795,012 + 818,506 + 859,407 + 905,508 + 925,828 + 921,091 + 900,389 = 8,584,449<br>50-59: 888,940 + 878,137 + 872,944 + 891,913 + 893,796 + 901,416 + 889,289 + 857,860 + 858,184 + 852,627 = 8,785,106<br>60+: 845,836 + 827,046 + 818,270 + 809,103 + 799,407 + 795,066 + 776,073 + 784,280 + 760,998 + 783,527 + 766,434 + 759,622 + 739,203 + 692,884 + 518,955 + 502,516 + 483,835 + 443,448 + 389,310 + 397,453 + 408,011 + 390,052 + 372,609 + 362,050 + 336,284 + 325,338 + 293,641 + 280,250 + 250,255 + 226,053 + 186,015 + 160,562 + 132,403 + 110,466 + 89,330 + 69,801 + 53,201 + 39,728 +29,030 + 20,035 + 21,860 = 17,850,240<br><br>Total population: 67,063,703 |
| Inbounding Travellers (China to France) (Dec 2022)<br><br>France!B8, France!B9 | https://www.entreprises.gouv.fr/files/files/directions_services/etudes-et-statistiques/Chiffres_cles/Tourisme/2019-04-key-facts-on-tourism-2018.pdf<br><br>https://www.connexionfrance.com/article/French-news/Tourism-in-France-The-2022-trends-and-outlook-for-2023<br><br>In 2018, of the 68.1 million travelers to France, 2.1 million were |

| | |
|---|---|
| | from China, for a proportion of 2.1/68.1 = 0.030837.<br><br>The World Travel and Tourism Council reported that France welcomed 34.5 million tourists in 2022. An estimated 34,500,000*0.030837 = 1,063,877 tourists are from China (2915 per day). |
| Outbounding Travellers (France to China) (2018)<br><br>France!B10 | http://www.stats.gov.cn/sj/ndsj/2019/indexeh.htm<br><br>The number of travelers from France to China was extracted from the China National Bureau of Statistics through the China Statistical Yearbook 2019 (17-13: Number of Oversea Visitor Arrivals by Country/Region).<br><br>Outbound Travelers: 499,600 |
| Outbounding Travellers (France to China) (2023)<br><br>France!B11, France!B12, France!B13 | Using the McKinsey estimate, we can scale the estimated number of outbound travelers from France to China.<br><br>Feb 2023 = 499,600*2,000,000/145,307,800 = 6,876<br>Daily = 6,876/28 = 245.6 |
| Length of Stay (China to France) (2021)<br><br>France!B14 | https://www.statista.com/statistics/1246755/length-of-stay-overseas-tourists-in-france/<br><br>The average length of stay for inbound tourists to France in 2018 was around 6.68 days. |
| Length of Stay (France to China) (2019)<br><br>France!B15 | https://www.wta-web.org/wp-content/uploads/2022/03/China-Inbound-Tourism-Development-Report.pdf<br><br>The World Tourism Alliance estimated the average length of stay for inbound tourists to China was around 9.2 days (page 49). |
| Cumulative Cases<br><br>France!B16 | https://covid19.who.int/region/euro/country/fr<br><br>The cumulative cases during the 21.5-month period of infection-derived immunity durability was calculated by subtracting the cumulative number of cases from Feb 19th, 2023, by the cumulative number of cases from May 1st, 2021.<br><br>***Methodology***<br>Select “Cumulative”. Select “Daily”. Hover over the desired data and choose the number for confirmed cases. |

| | |
|---|---|
| | May 1st, 2021: 5,205,996<br>Feb 19th, 2023: 38,483,144<br><br>Cumulative Cases: 38,483,144 - 5,205,996 = 33,277,148 |
| Weekly Cases<br><br>France!B17, France!B18 | https://covid19.who.int/region/euro/country/fr<br><br>The weekly cases was extracted from the official WHO database for the week of Feb 13th to Feb 19th.<br><br>***Methodology***<br>Select "Daily Change". Select "Weekly". Hover over Feb 13th, 2023, and choose the number for confirmed cases.<br><br>Weekly Cases: 23,862<br>Daily Incidence: 23,862/7 = 3,409 |

**Germany**

| Data | Link and Extraction Methods |
|---|---|
| Vaccine coverage: At least one dose<br><br>Germany!B2:H2 | https://ourworldindata.org/covid-vaccinations<br><br>The coverage for individuals who took at least one dose of the vaccine during the 20.5-month period of vaccine durability was calculated by subtracting the cumulative percentage from Feb 13th, 2023, by the cumulative percentage from Jun 1st, 2021. The maximum of the percentage from this calculation and the calculation from **Vaccine coverage: Full course** was used.<br><br>***Methodology***<br>Set the country as "Germany". Set Metric to "People vaccinated". Select "Relative to Population".<br><br>Jun 1st, 2021: 44.46%<br>Feb 13th, 2023: 77.81%<br><br>Vaccine coverage: 77.81% - 44.46% = 33.35% |
| Vaccine coverage: Full course<br><br>Germany!B3:H3 | https://ourworldindata.org/covid-vaccinations<br><br>The coverage for individuals who took the full course of the vaccine during the 20.5-month period of vaccine durability was calculated by subtracting the cumulative percentage from Feb 13th, 2023, by the cumulative percentage from Jun 1st, 2021. |

| | |
|---|---|
| | ***Methodology***<br>Set the country as "Germany". Set Metric to "People fully vaccinated". Select "Relative to Population".<br><br>Jun 1st, 2021: 18.94%<br>Feb 13th, 2023: 76.24%<br><br>Vaccine coverage: 76.24% - 18.94% = 57.3% |
| Infection-derived immunity<br><br>Germany!B4:H4 | Infection-derived immunity was calculated by dividing the number of cumulative cases over the last 21.5 months, over the duration of the durability of immunity, by the total population.<br><br>Infection-derived immunity: 34,612,784/83,237,124 = 0.4158335 |
| Prevalence<br><br>Germany!B5:H5 | Prevalence was calculated by dividing the weekly cases by the total population.<br><br>Prevalence: 97,597/83,237,124 = 0.0011725 |
| Population size<br><br>Germany!B6:H6 | https://www.destatis.de/EN/Themes/Society-Environment/Population/Current-Population/Tables/lrbev01ga.html<br><br>***Methodology***<br>*Notes*<br>The population is presented in percentages. The percentages are assumed to be equally distributed across each age in the chart. For example, the 20-39 age group is split evenly between 20-29 and 30-39.<br><br><18: 83,237,124*16.7% = 13,900,600<br>18-29: 83,237,124*(18.5% - 16.7% + 24.4%/2) = 7,075,156<br>30-39: 83,237,124*(24.4%/2) = 10,154,929<br>40-49: 83,237,124*(27.7%/2) = 11,528,342<br>50-59: 83,237,124*(27.7%/2) = 11,528,342<br>60+: 83,237,124*(22.0% + 7.3%) = 24,388,477<br><br>Total population: 83,237,124 |
| Inbounding Travellers (China to Germany) (Dec 2022)<br><br>Germany!B8, Germany!B9 | https://www.destatis.de/DE/Themen/Branchen-Unternehmen/Gastgewerbe-Tourismus/Publikationen/Downloads-Tourismus/statistischer-bericht-monatserhebung-tourismus-2060710231025.html<br><br>Germany's Statistics Department reports that 22,063 people visited Germany in Feb 2023 from China (788 per day). |
| Outbounding Travellers | http://www.stats.gov.cn/sj/ndsj/2019/indexeh.htm |

| | |
|---|---|
| (Germany to China) (2018)<br><br>Germany!B10 | The number of travelers from Germany to China was extracted from the China National Bureau of Statistics through the China Statistical Yearbook 2019 (17-13: Number of Oversea Visitor Arrivals by Country/Region).<br><br>Outbound Travelers: 643,700 |
| Outbounding Travellers (Germany to China) (2023)<br><br>Germany!B11, Germany!B12, Germany!B13 | Using the McKinsey estimate, we can scale the estimated number of outbound travelers from Germany to China.<br><br>Feb 2023 = 643,700*2,000,000/145,307,800 = 8,860<br>Daily = 8,860/28 = 316.4 |
| Length of Stay (China to Germany) (2021)<br><br>Germany!B14 | https://www.statista.com/statistics/572316/trip-duration-german-tourists/<br><br>The average trip in Germany was 12.7 days long in 2022. |
| Length of Stay (Germany to China) (2019)<br><br>Germany!B15 | https://www.wta-web.org/wp-content/uploads/2022/03/China-Inbound-Tourism-Development-Report.pdf<br><br>The World Tourism Alliance estimated the average length of stay for inbound tourists to China was around 9.2 days (page 49). |
| Cumulative Cases<br><br>Germany!B16 | https://covid19.who.int/region/wpro/country/kr<br><br>The cumulative cases during the 21.5-month period of infection-derived immunity durability was calculated by subtracting the cumulative number of cases from Feb 19th, 2023, by the cumulative number of cases from May 1st, 2021.<br><br>***Methodology***<br>Select "Cumulative". Select "Daily". Hover over the desired data and choose the number for confirmed cases.<br><br>May 1st, 2021: 3,402,021<br>Feb 19th, 2023: 38,014,805<br><br>Cumulative Cases: 38,014,805 - 3,402,021 = 34,612,784 |
| Weekly Cases<br><br>Germany!B17, Germany!B18 | The number of weekly cases was extracted from the official WHO database for the week of Feb 13th to Feb 19th.<br><br>***Methodology*** |

| | |
|---|---|
| | Select “Daily Change”. Select “Weekly”. Hover over Feb 13th, 2023, and choose the number for confirmed cases.<br><br>Weekly Cases: 97,597<br>Daily Incidence: 97,597/7 = 13,942 |

**Italy**

| Data | Link and Extraction Methods |
|---|---|
| Vaccine coverage: At least one dose<br><br>Italy!B2:H2 | https://ourworldindata.org/covid-vaccinations<br><br>The coverage for individuals who took at least one dose of the vaccine during the 20.5-month period of vaccine durability was calculated by subtracting the cumulative percentage from Feb 13th, 2023, by the cumulative percentage from Jun 1st, 2021. The maximum of the percentage from this calculation and the calculation from **Vaccine coverage: Full course** was used.<br><br>***Methodology***<br>Set the country as “Italy”. Set Metric to “People vaccinated”. Select “Relative to Population”. Set Interval to “Cumulative”<br><br>Jun 1st, 2021: 40.96%<br>Feb 13th, 2023: 86.23%<br><br>Vaccine coverage: 86.23% - 40.96% = 45.27% |
| Vaccine coverage: Full course<br><br>Italy!B3:H3 | https://ourworldindata.org/covid-vaccinations<br><br>The coverage for individuals who took the full course of the vaccine during the 20.5-month period of vaccine durability was calculated by subtracting the cumulative percentage from Feb 13th, 2023, by the cumulative percentage from Jun 1st, 2021.<br><br>***Methodology***<br>Set the country as “Italy”. Set Metric to “People fully vaccinated”. Select “Relative to Population”.<br><br>Jun 1st, 2021: 21.13%<br>Feb 13th, 2023: 81.24%<br><br>Vaccine coverage: 81.24% - 21.13% = 60.01% |
| Infection-derived immunity | Infection-derived immunity was calculated by dividing the number of cumulative cases over the last 21.5 months, over the duration of |

| | |
|---|---|
| Italy!B4:H4 | the durability of immunity, by the total population.<br><br>Infection-derived immunity: 21,533,330/58,870,750 = 0.3657730 |
| Prevalence<br><br>Italy!B5:H5 | Prevalence was calculated by dividing the weekly cases by the total population.<br><br>Prevalence: 4,164/58,870,750 = 0.0000707 |
| Population size<br><br>Italy!B6:H6 | https://www.populationpyramid.net/italy/2023/<br><br>The population sizes for each age group were extracted from PopulationPyramid.net, which uses information from the United Nations.<br><br>***Methodology***<br>*Notes*<br>The age group of 15-19 was separated across the <18 and 18-29 groups by assuming an equal number of individuals for each age. Females and males for each age group were added together.<br><br><18: 1,062,733 + 1,005,264 + 1,227,328 + 1,160,645 + 1,402,208 + 1,323,106 + (1,470,662 + 1,384,622)*3/5 = 8,894,454<br>18-29: (1,470,662 + 1,384,622)*2/5 + 1,520,070 + 1,402,357 + 1,561,592 + 1,438,384 = 7,064,517<br>30-39: 1,642,757 + 1,576,820 + 1,686,485 + 1,660,975 = 6,567,037<br>40-49: 1,857,515 + 1,852,236 + 2,201,241 + 2,221,798 = 8,132,790<br>50-59: 2,355,735 + 2,409,334 + 2,364,241 + 2,455,115 = 9,584,425<br>60+: 2,041,653 + 2,185,911 + 1,733,590 + 1,912,112 + 1,537,518 + 1,753,972 + 1,281,472 + 1,552,547 + 953,762 + 1,306,348 + 561,194 + 919,755 + 216,148 + 470,606 + 43,050 + 136,006 + 3,894 + 17,989 = 18,627,527<br><br>Total population: 58,870,750 |
| Inbounding Travellers (China to Italy) (Feb 2023)<br><br>Italy!B8, Italy!B9 | https://stats.oecd.org/index.aspx?DataSetCode=TOURISM_INBOUND<br><br>The OECD documents that in 2021, of the 26,903,217 overnight visitors to Italy, 204,874 were from China, or roughly a proportion of 204,874/26,903,217 = 0.00762<br><br>The Bank of Italy reports that in February 2023, 4,751,000 people visited Italy. The number of Chinese travelers can be estimated to be 4,751,000*0.00762 = 36,180 (1,292 per day) |

| | |
|---|---|
| Outbounding Travellers (Italy to China) (2018)<br><br>Italy!B10 | http://www.stats.gov.cn/sj/ndsj/2019/indexeh.htm<br><br>The number of travelers from Italy to China was extracted from the China National Bureau of Statistics through the China Statistical Yearbook 2019 (17-13: Number of Oversea Visitor Arrivals by Country/Region).<br><br>Outbound Travelers: 278,100 |
| Outbounding Travellers (Italy to China) (2023)<br><br>Italy!B11, Italy!B12, | Using the McKinsey estimate, we can scale the estimated number of outbound travelers from Italy to China.<br><br>Feb 2023 = 278,100*2,000,000/145,307,800 = 3,828<br>Daily = 3,828/28 = 136.7 |
| Length of Stay (China to Italy) (2021)<br><br>Italy!B13 | https://www.statista.com/statistics/901296/number-of-nights-spent-by-chinese-tourists-in-accommodations-in-italy/<br><br>Chinese tourists spent an approximate 5.36 days in Italy in 2019. |
| Length of Stay (Italy to China) (2019)<br><br>Italy!B14 | https://www.wta-web.org/wp-content/uploads/2022/03/China-Inbound-Tourism-Development-Report.pdf<br><br>The World Tourism Alliance estimated the average length of stay Italian visitors to China was around 7.2 days (page 96). |
| Cumulative Cases<br><br>Italy!B15 | https://covid19.who.int/region/euro/country/it<br><br>The cumulative cases during the 21.5-month period of infection-derived immunity durability was calculated by subtracting the cumulative number of cases from Feb 19th, 2023, by the cumulative number of cases from May 1st, 2021.<br><br>***Methodology***<br>Select “Cumulative”. Select “Daily”. Hover over the desired data and choose the number for confirmed cases.<br><br>May 1st, 2021: 4,022,653<br>Feb 19th, 2023: 25,555,983<br><br>Cumulative Cases: 25,555,983 - 4,022,653 = 21,533,330 |
| Weekly Cases<br><br>Italy!B16, Italy!B17 | https://covid19.who.int/region/euro/country/it<br><br>The weekly cases was extracted from the official WHO database for the week of Feb 13th to Feb 19th. |

| | |
|---|---|
| | ***Methodology***<br>Select “Daily Change”. Select “Weekly”. Hover over Feb 13th, 2023, and choose the number for confirmed cases.<br><br>Weekly Cases: 29,146<br>Daily Incidence: 29,146/7 = 4,164 |
